# Extracting TNFi Switching Reasons and Trajectories From Real-World Data Using Large Language Models

**DOI:** 10.1101/2025.04.14.25325834

**Authors:** Brenda Y Miao, Marie Binvignat, Augusto Garcia-Agundez, Maxim Bravo, Christopher YK Williams, Claire Q Miao, Ahmed Alaa, Vivek Rudrapatna, Atul J Butte, Gabriela Schmajuk, Jinoos Yazdany

## Abstract

**Importance:** Tumor necrosis factor inhibitors (TNFi) are widely used for auto-immune conditions. Despite their efficacy, many patients switch TNFis due to lack of efficacy, cost-related reasons, or adverse events. Understanding why switches occur is important, but requires extensive chart review.

**Objective:** To determine whether large language models (LLMs) can automatically perform chart review, accurately identifying TNFi switching trajectories and reasons for switching in a large real-world cohort.

**Design:** Observational study using de-identified electronic health record data (2012-2023). Medication orders and associated clinical notes for TNFi agents were extracted; at least 6 months of follow-up was required to ascertain switches.

**Setting:** Single academic medical center (University of California, San Francisco).

**Participants:** 9,187 patients (mean [SD] age, 39.9 [19.0] years; 57.1% female) who received ≥1 TNFi with adequate follow-up. Among these, 1,481 (16.1%) had ≥1 TNFi switch, 418 (4.5%) had ≥2 switches, and 150 (1.6%) had ≥3 switches.

**Exposures:** Switching was defined as a change from one TNFi to a different TNFi at consecutive encounters.

**Main Outcome(s) and Measure(s):** Using GPT-4, we extracted which TNFi was stopped or started, and reasons for switching: adverse event; drug resistance; insurance/cost; lack of efficacy; patient preference; other; unknown. Performance was compared with eight open source LLMs, structured medication data and expert annotations.

**Results:** After applying inclusion criteria, 3,104 switches between different TNFi drugs in 2,112 patients were identified. GPT-4 achieved micro-F1 scores of 0.75 for stopped TNFi, 0.80 for started TNFi, and 0.83 for switch reason. From all open-source models, Starling-7B-beta and Llama-3-8B offered the most competitive performance overall compared to GPT-4 and achieved similar win-loss ratios. The primary reasons identified by GPT-4 was lack of efficacy (56.9%), followed by adverse events (13.5%) and insurance/cost (10.8%).

**Conclusions and Relevance:** Both GPT-4 and locally deployable LLMs, demonstrated potential in executing complex reasoning tasks, specifically identifying reasons for switching between TNF inhibitors. This finding suggests broader application in clinical research and documentation. Further research is needed to assess model performance across additional medication classes and patient populations.

**Keys points:** *Question:* Can large language models (LLMs) identify (TNF inhibitors) TNFi switching trajectories and reasons from clinical notes?

*Findings:* We used de-identified electronic health records from UCSF (University of California San Francisco) from 9,187 patients who received ≥1 TNFi. GPT-4 achieved micro-F1 scores up to 0.830 identifying reasons and specific TNFi starts/stops compared to clinical expert annotations, surpassing eight open-source LLMs. The best open-source models, Llama-3-8b-chat-hf and Starling-7B-beta, matched GPT-4 in determining which TNFi was started but had lower accuracy in identifying reasons for switching.

*Meaning:* LLMs evaluated in this study were capable of performing complex reasoning tasks in identifying reasons for switching between TNFi. Broader application could be used for other biological but also in other pharmacoepidemiology studies and in chart summarization.

## Introduction

Tumor necrosis factor inhibitors (TNFi) are a class of biologic therapies indicated for managing multiple autoimmune diseases, including Inflammatory Bowel Disease^1,2^ (IBD) and Rheumatoid Arthritis^3,4^ (RA). While there are now many TNFi and biosimilars available, there are few biomarkers or clinical recommendations to guide therapeutic selection of specific medications within this drug class for individual patients. It is therefore not surprising that the initial choice of agent varies among physicians. ^3,5^

Previous studies indicate that approximately 14.5% of patients with IBD switch medications at least once, primarily to another TNFi.^6^ In a cohort of US patients with RA, 39.3% who did not respond to a first-line TNFi switched to another TNFi.^7^ There are several additional reasons that could explain switches between TNFi. Some patients may develop anti-TNFi antibodies, leading to a loss of drug efficacy.^2,8^ Insurance coverage policies may also lead to switching. While several risk factors have been associated with TNFi switching, such as female sex, age, or high disease activity^5,9^, patient switch trajectories are not easily predictable. In addition, a high TNFi switch rate is associated with poorer disease control and higher annual treatment costs^10^.

Most studies of switching between different TNFi are based on structured analyses of medical record data or clinical trial results, and can overlook potential reasons for switching that may not be available in these data sources but may be discussed in clinical notes, such as insurance issues, patient preferences, adverse events, and other real-world factors influencing treatment changes.^11^ Clinical notes are an underutilized resource to elicit reasons for medication switching because they require time-consuming manual clinical review, making their use prohibitively long and costly.

In this context, Large Language Models (LLMs) such as GPT-4 may offer a promising, cost-effective approach to automatically extract information from clinical notes, such as reasons for medication switching. Related work in other contexts shows that LLMs are capable of accurate clinical information extraction.^12,13^ In this study, we aim to profile patterns of switching between different TNFi and to use LLMs to understand the reasons for these switches. We also aim to perform a comparative analysis of different open-source LLMs on this task.

## Methods

### TNFi cohort selection

We identified a **TNFi**-treated patient cohort using the University of California, San Francisco (UCSF) Information Commons dataset, which contains longitudinal, de-identified medical record data and clinical notes between 2012 and 2023.^14^ We selected all TNFi medication orders and administrations, using a string search of all TNFi generic or brand names derived from data provided by the Food and Drug Administration.^15^ Medication names were mapped to appropriate generic or biosimilar categories (**Table S1**), and encounters where a patient switched from one TNFi to another, defined as a change in TNFi prescribed between two consecutive encounters, and had an associated clinical note were identified. Patients without any clinical and demographic data (including age, sex, race, ethnicity) were excluded. We also excluded patients who did not have an encounter at least 6 months following the medication order, since in cases with insufficient follow up time it could not be determined whether the patient switched TNFi in these cases. We excluded, for consistency purposes, encounters where multiple TNFi were ordered on the same date. In addition, for encounters with multiple notes, only the first note was used for analysis. Diagnosis codes linked to TNFi prescriptions were manually reviewed by a physician (MB) and categorized into the following groups: IBD, Psoriatic Arthritis, Juvenile Idiopathic Arthritis (JIA), Rheumatoid Arthritis (RA), Spondyloarthritis (SA), Uveitis, Hidradenitis suppurativa, Sarcoidosis, Vasculitis, and Unspecified. Disease categories with fewer than 10 individuals who switched were consolidated into “Other” in accordance with de-identification guidelines. Patient demographic information was tabulated using the tableone package,^16^ with continuous distributions reported as means and standard deviations, and categorical values represented as proportions. All data used in this study was performed using deidentified data and was thus determined to be exempt from further review by the UCSF IRB.

### Prompt selection for TNFi switching reason extraction using GPT-4

The GPT-4 LLM was used to extract information about TNFi switching from associated clinical notes in a zero-shot manner (i.e. without dedicated training or examples of the task being provided). Data were split into 5%/95% validation and test sets, with the validation set used for evaluation of 4 different prompts (**Table S2**) and final metrics reported on the test set. Each prompt was used to extract the new TNFi started, the previous TNFi stopped, and one of the following categories for switching: adverse event, lack of efficacy, insurance/cost, drug resistance (defined by the documentation of anti-drug antibodies), patient preference, other, or unknown (“NA”). An example of a prompt used (“Reasons provided” prompt) is as follows:

> *“Task: Tumor necrosis factor inhibitors (TNFis) are biologic drugs targeting TNF proteins. Using the clinical note provided, extract the following information into this JSON format: {“new_TNFi“:“What new TNFi was prescribed or started? If the patient is not starting a new TNFi, write “NA“”,“last_TNFi“:“What was the last TNFi the patient used? If none, write “NA“”,“Reason for Switching“:“Which best describes why the last TNFi was stopped or planned to be stopped? “Adverse event”, “Drug resistance”, “Insurance/Cost”,“Lack of efficacy”,“Patient preference”,“Other”, “NA“”,“full_reason_last_TNFi_stopped“:“Provide a description for why the last TNFi was stopped or planned to be stopped?“} Answer:”*

Examples of extracted reasons are presented in **Table S3**. Model performance was assessed against silver-standard labels from associated medication order information as well as gold-standard annotations provided by a physician (MB) according to annotation guidelines (**Figure S1**) and microF1 scores were assessed. For silver-standard labels, we reported separate microF1 scores for all notes and only notes determined by GPT-4 to contain medication information. This aligns with previous studies showing that silver-standard labels derived from medication order data are a more reliable proxy of information extraction performance when used to assess for clinical notes containing medication information^17^. The prompt with the highest microF1 scores calculated using gold-standard annotations was used to extract TNFi switching information from the test set for downstream analysis.

### Comparison of open-source LLMs on TNFi switching information extraction

Several open-source models were also assessed using the manually annotated validation data. These included three models trained from scratch (“Yi-6B-Chat”, “Llama-2-7B-Chat”, “Gemma-7B-IT”), as well as updated versions (“Llama-3-8B-Instruct”) or further fine-tuned versions of some models (“zephyr-7b-gemma-v0.1”, “OpenHermes-2.5-Mistral-7B”, “Snorkel-Mistral-PairRM-DPO”, “Starling-7B-alpha”, “Starling-7B-beta”). Two models, “JSL-MedMNX-7B-SFT” and “BioMistral-7B” were specifically trained or fine-tuned on biomedical data. Additional details on models and parameters usage can be found in the supplemental figures (**Table S4**). Open source models were compared using microF1 scores, as well as average pairwise win rates compared to GPT-4 for each response, following comparative open-source benchmarks.^18^ The pairwise win rate is an assessment of how often a model provides a correct response when another model does not. Model “ties” were recorded when both models provide correct or incorrect values. We report mean win rates of each model against all other models.

## Results

## TNFi switching cohort from UCSF Information Commons

We identified 190,518 relevant TNFi medication orders (**Figure 1**), from 12,442 unique patients. These orders were mapped to generic names, ignoring dosage information and modality (**Table S1**). After removing 14 patients without demographic information, 190,500 total medication orders remained. Multiple and duplicate TNFi orders and orders without associated clinical notes were dropped, leaving 64,983 unique medication orders. When there were different TNFi order notes at the same encounter, only the first associated clinical note was considered for downstream analysis (often considered as the most informative). This left a TNFi treatment dataset consisting of 58,323 medication orders from 11,572 patients. Of these patients, 2,112 (18.25%) had a documented TNFi switch, while 7,075 (61.14%) had no documented switch with a follow-up encounter at least 6 months after the TNFi order. The remaining 2,385 (20.61%) patients also did not have a medication switch but were lost to follow-up and were excluded from further analysis.

**Figure 1.**
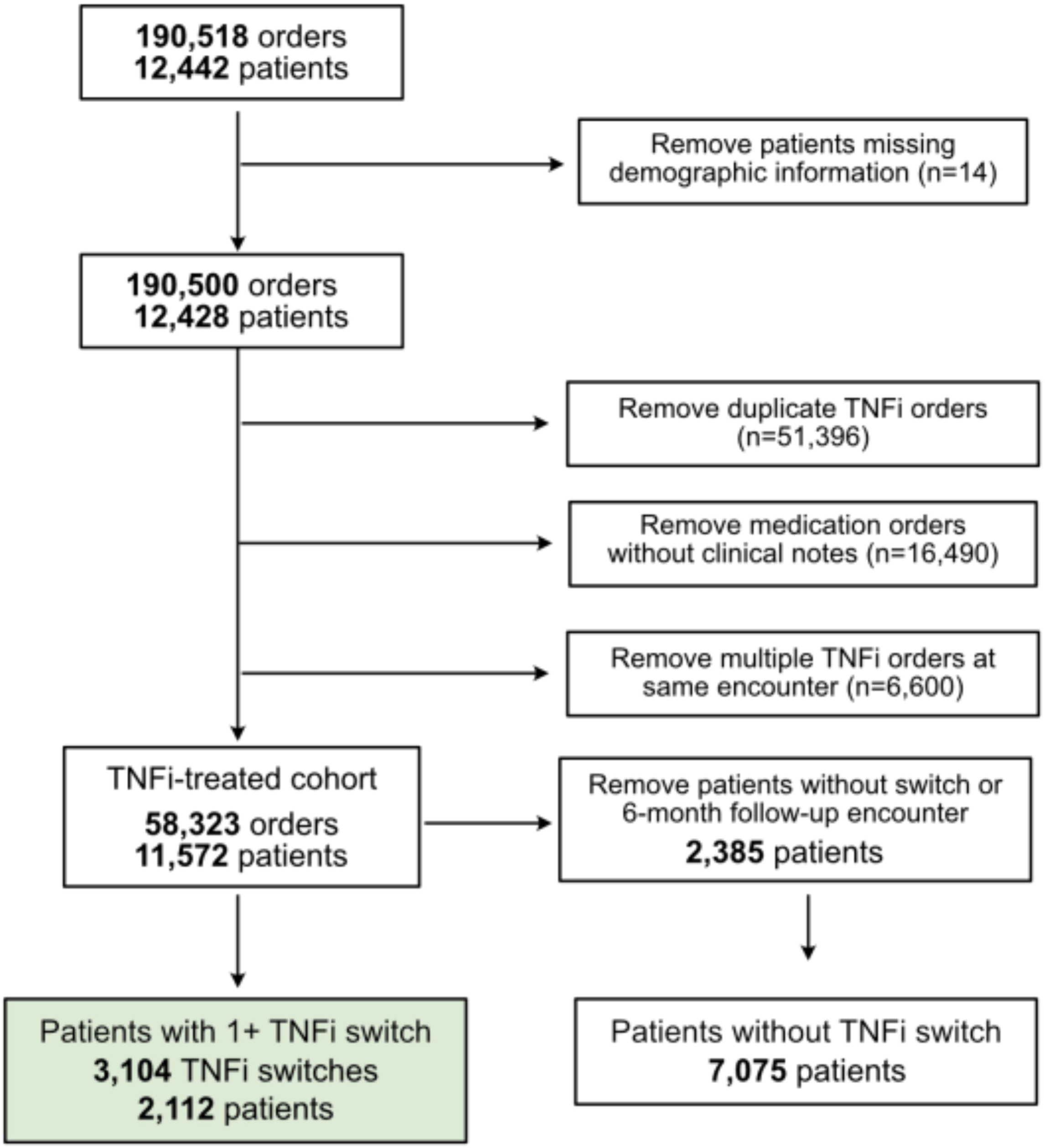
Flow chart and cohort selection. We identified 190,518 TNFi medication orders from 12,442 unique patients in the UCSF Information Commons dataset. After excluding 14 patients without demographic information, 190,500 orders from 12,428 patients remained. We further removed 51,396 duplicate TNFi orders, 16,490 orders without associated clinical notes, and 6,600 orders where multiple TNFi medications were ordered at the same encounter. The resulting TNFi-treated cohort consisted of 58,323 orders from 11,572 patients. Among these patients, 2,112 (18.3%) experienced at least one TNFi switch, representing 3,104 total switches.

### TNFi cohort demographics

The TNFi treatment cohort (n=9,187) had a mean age of 39.9 years (SD 19.0), with a slightly higher proportion of female patients (57.1%) (**Table 1**). Regarding self-reported race and ethnicity, the cohort was composed of 60.2% White, 13.9% Latinx, 7.0% Asian, and 4.8% Black or African American individuals. For the majority of TNFi orders, associated disease was not filled out from medication switching order and structured data. Within known diseases, the most common primary diagnosis was inflammatory bowel disease (IBD) (15.4%), followed by rheumatoid arthritis (RA) (5.7%) and juvenile idiopathic arthritis (JIA) (3.1%). Additional diagnoses included psoriasis, spondyloarthritis, and uveitis. A subset of patients (7.4%) had multiple diagnoses. The mean follow-up time was 5.7 years (SD 4.9). The most common first TNFi across all patients was adalimumab (40.9%). Infliximab (26.3%) and etanercept (24.1%) were the next most prescribed.

**Table 1.**
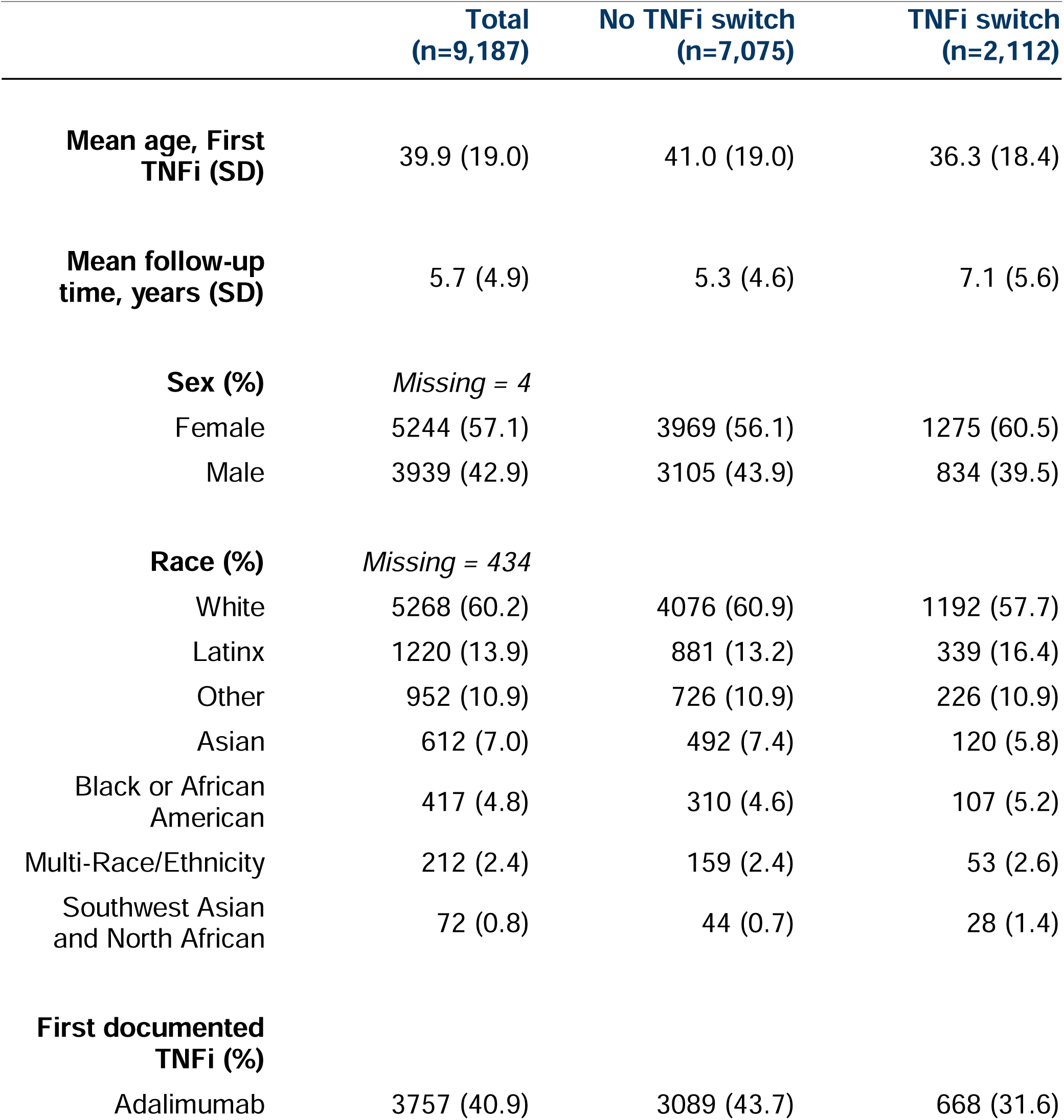
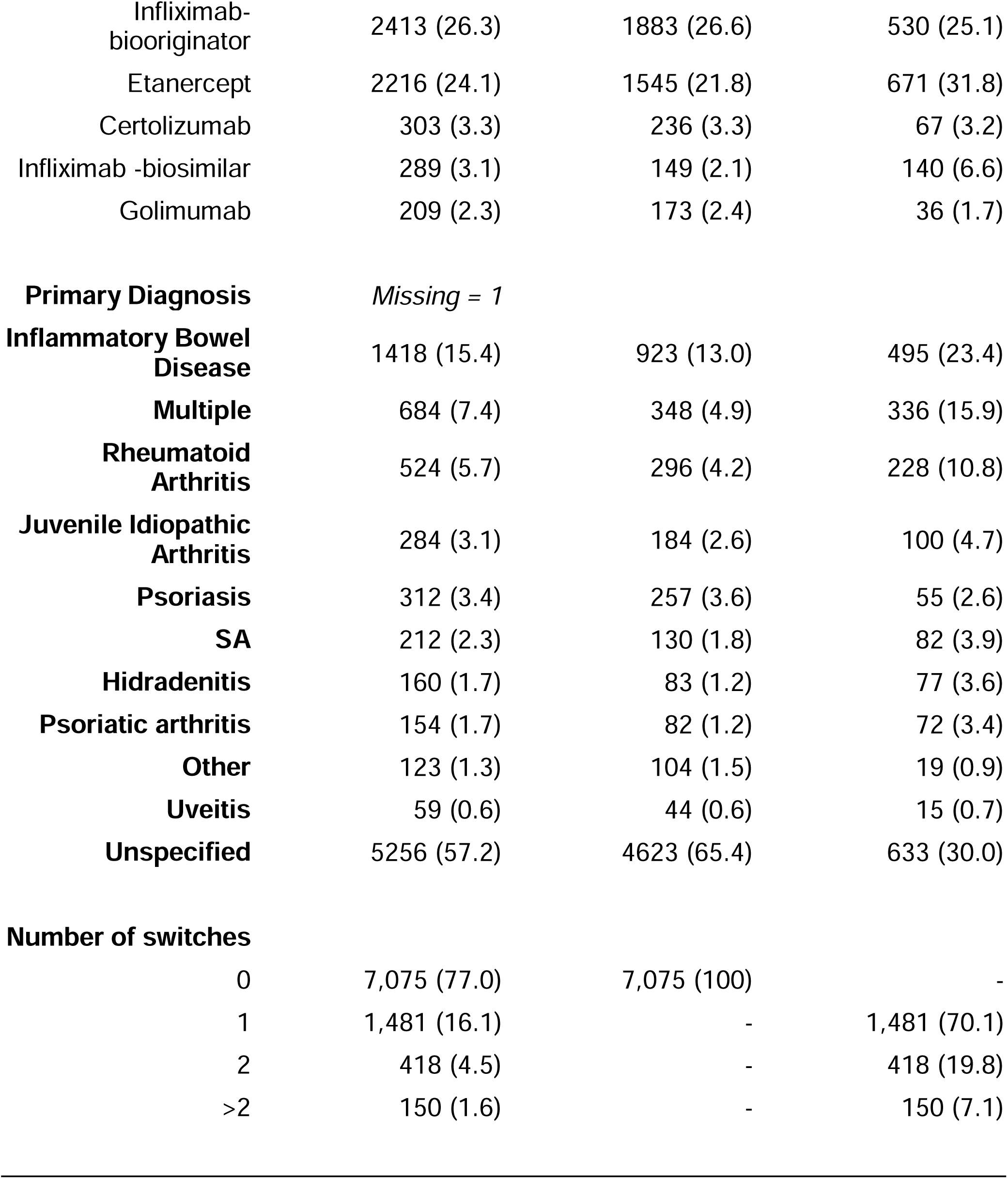
Patient Demographics for the TNFi Cohort Extracted from UCSF Information Commons. Patient demographics, including age, sex, race and ethnicity, mean follow-up duration, and initial documented TNFi and disease, are presented for 9,187 patients in the UCSF TNFi cohort. Data are stratified based on switching between different TNFi (no switch vs. any switch).

Within this cohort, 1,481 patients (16.1%) had at least one TNFi switch. Among these, 418 patients (19.8%) had two switches, and 150 patients (7.1%) had three or more switches (**Figure 2, Table S5**).

**Figure 2.**
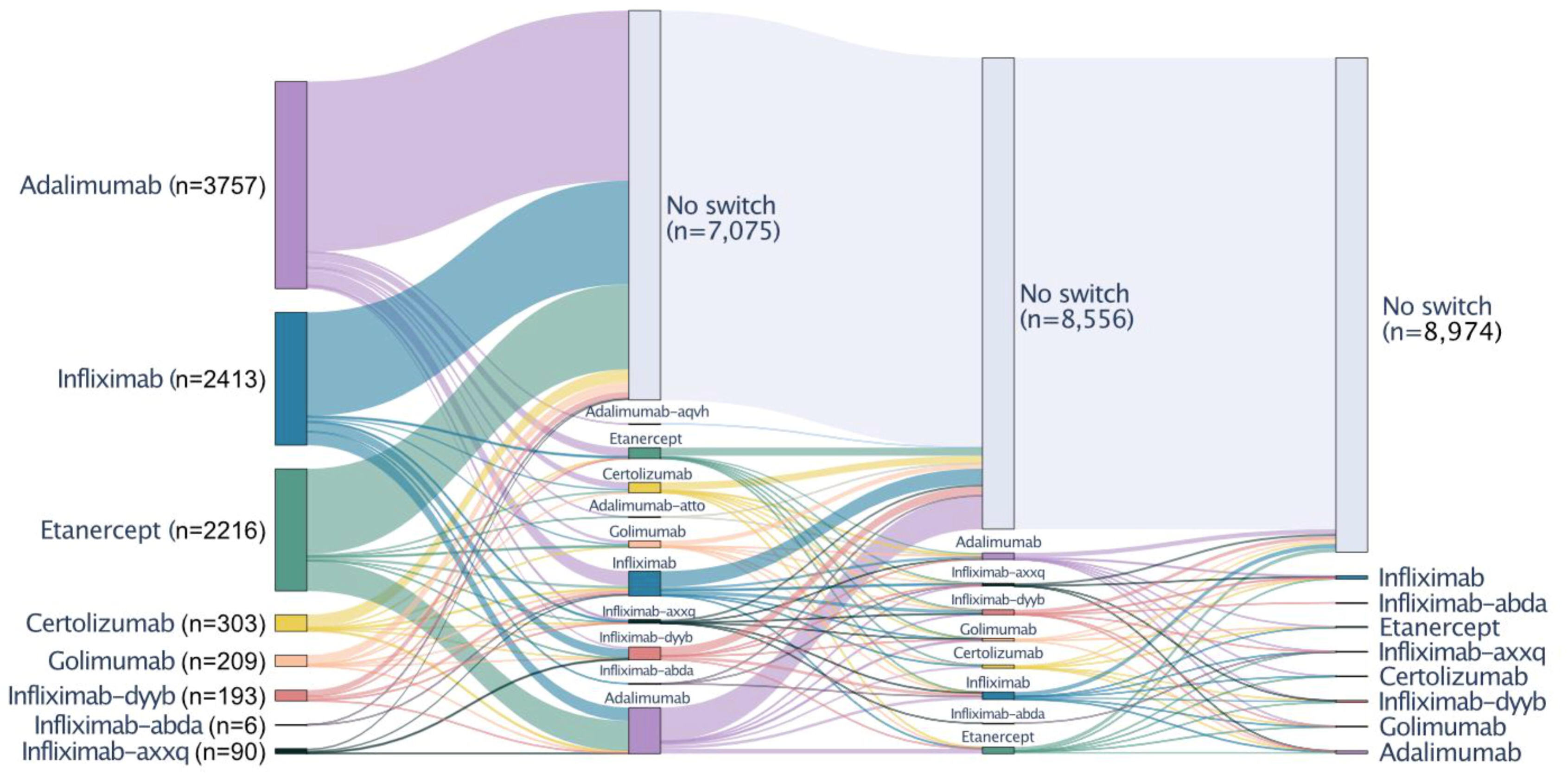
Sankey diagram for TNFi switching trajectories. TNFi treatment patterns among 9,187 patients identified from UCSF information commons through the *GPT-4-turbo-128k* (“GPT-4”) model. The diagram displays the relative proportion of patients switching or not switching TNFi.

### Prompt development

The GPT-4-turbo-128k (“GPT-4”) model was used to test four different prompts for extracting information about TNFi switching strategies and reasons for switching (**Table S2**). Out of the default prompt, prompt that provided specific categories for names of TNFi to extract (“Drugs provided”), a prompt that specified categorical reasons for switching (“Reasons provided”), or both drugs and reason categories provided (“All values provided”), the prompt providing the reason categories had the best overall performance (**Figure 3A**). With this prompt, microF1 scores were 0.42 for TNFi stopping information extraction and 0.50 for extracting which new TNF was prescribed (n=146). When excluding notes that did not contain a reason for a switch, as determined by GPT-4, microF1 scores increased to 0.63 (n=71) and 0.89 (n=56), respectively for TNFi stopping and new TNFi order information (**Table S6**). This was comparable to GPT-4 performance assessed against gold-standard clinical annotations, which showed microF1 scores up to 0.75 and 0.80, for TNFi stopped and started, respectively (**Table S7**). Although performance across all prompts were all within 0.05 by microF1 score, the “Reasons provided” prompt had the highest average microF1 score across all tasks and was used for all downstream tasks.

**Figure 3.**
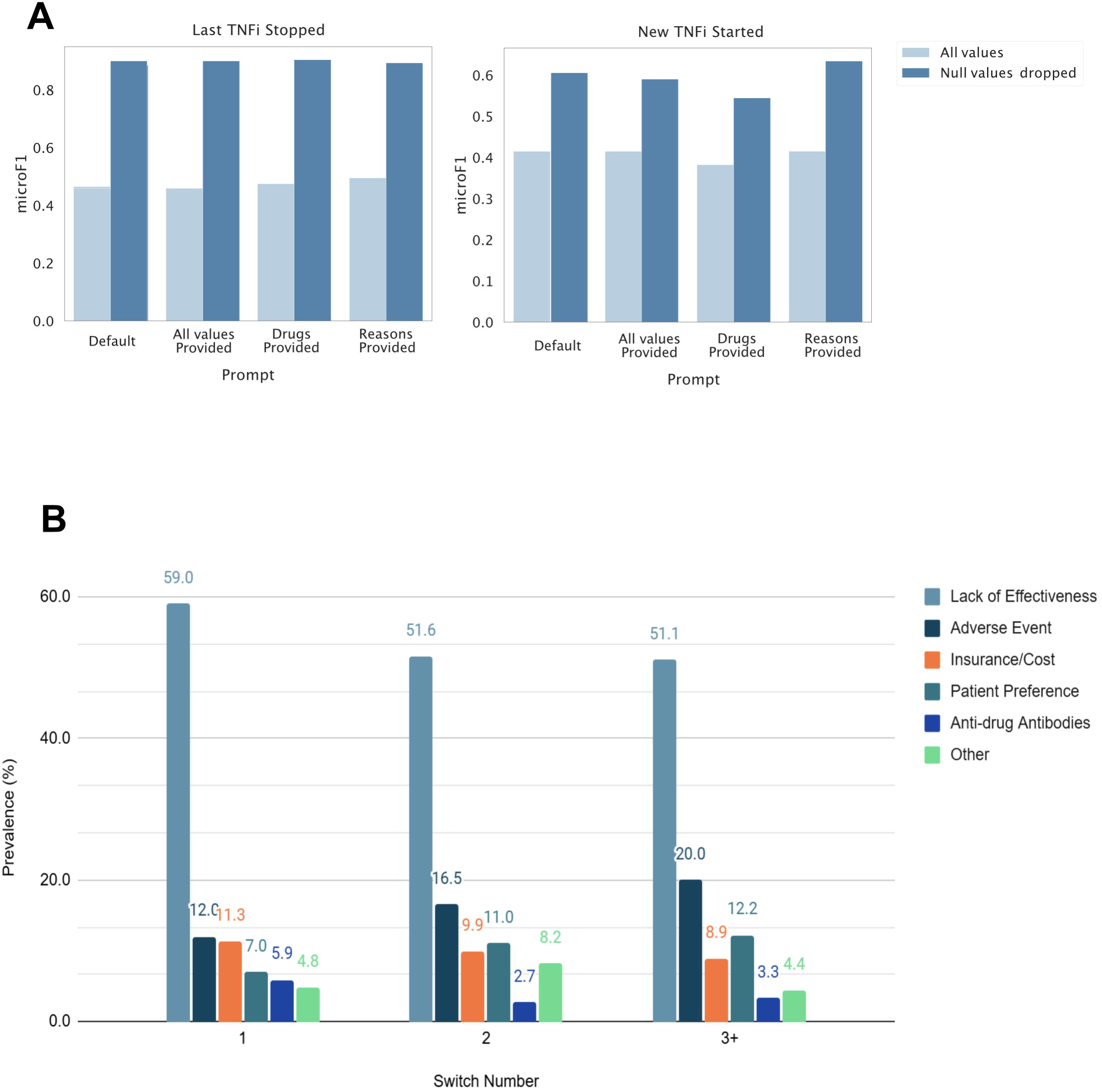
**A. Automated evaluation of GPT-4-turbo-128k performance across different prompts. GPT-4-turbo-128k was used to extract TNFi switching information,** including which TNFi was stopped and which was started. microF1 scores evaluated against structured data are shown, with LLM extracted null values included (“All values”) and counted as incorrect, or when excluding notes that did not contain a reason for switching (“Null values dropped”). **B. Reason for TNFi switching bar plot from GPT-4-turbo-128k model.** Proportion of reasons for TNFi switching in UCSF information commons extracted by GPT-4-turbo-128k, categorized based on the sequences of switches in the patient’s trajectory (first, second, and third or more switches). Identified reasons for switching included adverse events, drug resistance (defined as documented presence of anti-drug antibodies), insurance/cost, lack of efficacy, patient preference or other.

### Reasons for TNFi switching using GPT-4 abstracted information

When the best prompt (“Reasons provided”) was applied to the test dataset (n=2958), GPT-4 performance on TNFi started and stopped information extraction, compared to structured medication order information, demonstrated microF1 scores of 0.51 and 0.37, respectively. Analysis of all the reasons for TNFi switching extracted by GPT-4 for the validation and test sets uncovered that 1759 (59%) of the notes appeared to contain no reasons for switching. When excluding these notes, microF1 scores increased to 0.90 (n=1184) and 0.60 (n=1331), respectively. The most commonly extracted reason for switching was lack of efficacy (n=568, 56.9%) and adverse events (n=135, 13.5%). Insurance or cost issues accounted for 10.8% (n=108) of the TNFi switches and patient preference for another 8.2% (n=82) (**Table S8, Figure 3B**).

### Comparison of TNFi information extraction across LLMs

The best prompt previously selected (“Reasons provided”) was also used to understand how different open source LLMs (**Figure 4, Figure S2**) performed on these treatment information extraction tasks compared to GPT-4. MicroF1 scores of this prompt across open-source models showed that the “Starling-7b-beta” model had the highest average microF1 score of 0.52 while “Llama-2-7B-chat” had the lowest average score of 0.07 (**Table S3**). Again, only evaluating notes that contained reasons for TNFi switching increased microF1 scores, which ranged from 0.42 for “Llama-2-7B-chat” to 0.85 for “Starling-7b-beta”. When compared to gold-standard annotations, “starling-7b-alpha” had the highest F1 score in which previous TNFi was stopped (0.897), while GPT-4 had the highest F1 scores identifying which TNFi was started (0.801) and reasons for switching (0.825) (**Table S5**).

**Figure 4.**
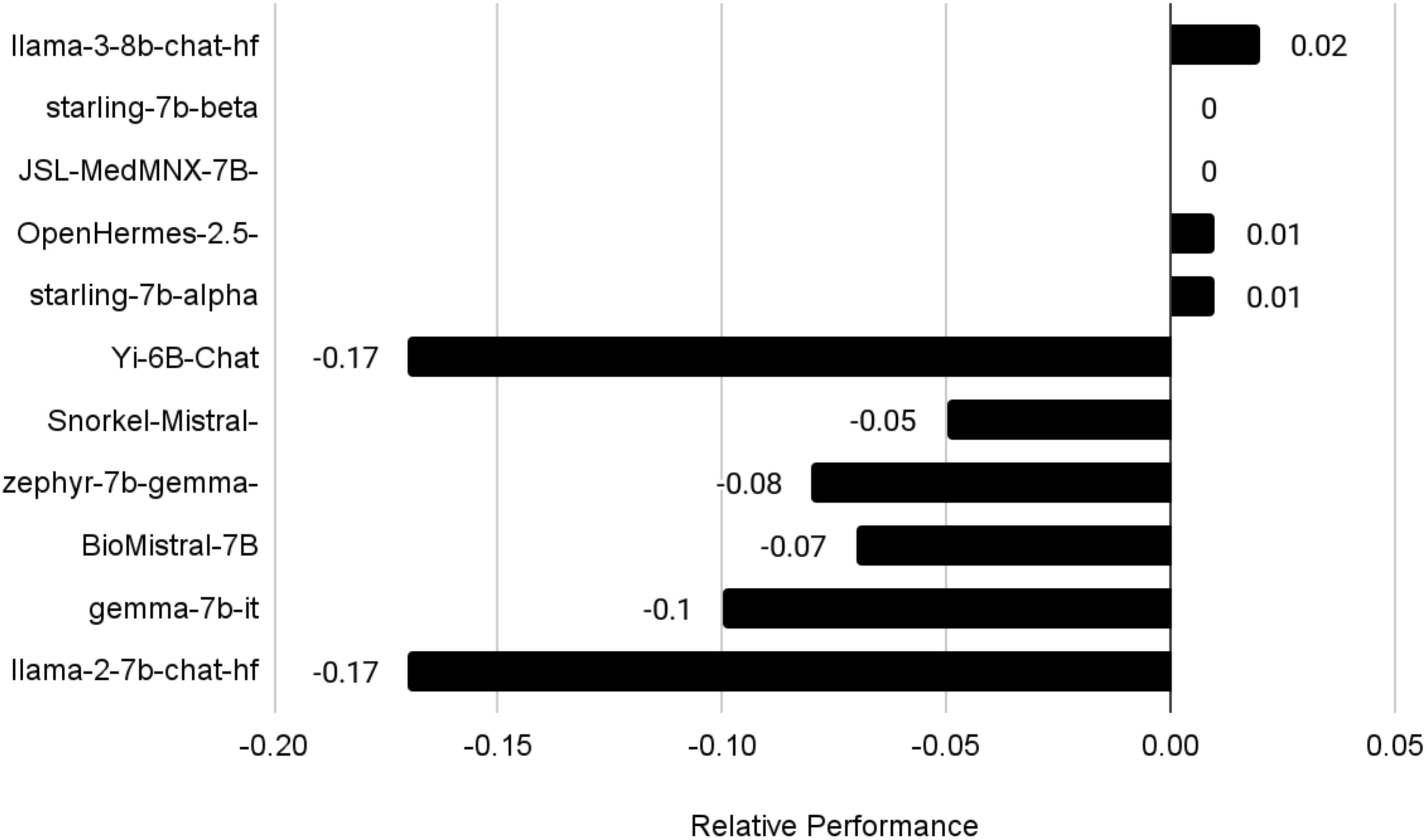
Average Win minus Loss Rates of Large Language Models Compared to GPT-4. Positive numbers indicate the model is superior to GPT-4 (win rate is higher than loss rate), negative numbers indicate the contrary. A complete heatmap with all model comparisons is available in Figure S4.

The concordance between models was also explored. Given GPT-4 performance on the previous tasks, outputs from this model were used as a baseline to further evaluate pairwise concordance between other models (**Table S9**). Llama-3-8B-Instruct and Starling-7B-beta showed the highest mean concordance with GPT-4 extracted information, with concordance rates of 82.4% (SD: 0.6%) and 77.0% (SD: 6.0%), respectively. Llama-2-7b-Chat showed the lowest concordance, with only 55.3% of values concordant (SD: 7.6%). We also evaluated pairwise win and tie rates of these models compared to GPT-4 extracted values. Mean tie rates ranged from 66.3% (SD: 16.2%) for llama-2-7b-chat-hf to 80.0% (SD: 12.8%) for JSL-MedMNX-7B-SFT. Llama-3-8B-Instruct had the highest average win rate at 15.5% (SD: 11.4%), followed by zephyr-7b-gemma-v01 at 12.7% (SD: 9.0%).

## Discussion

This study demonstrates the feasibility of using LLMs to extract TNFi switching reasons from clinical notes, offering insights into the predominant reasons for switching across a diverse population of patients. Our findings reveal that lack of efficacy, followed by adverse events, and insurance or cost considerations are among the most frequently documented reasons for TNFi switching. These results align with prior literature indicating that treatment failure and immunogenicity drive many TNFi switches.^4,6^

In our comparisons of proprietary and open-source LLMs, several open-source models performed nearly as well as GPT-4 in extracting TNFi switches (**Figure 5**) but displayed greater variability in identifying specific reasons for switching. For example, some models achieved higher F1 scores in identifying the previous TNFi or the newly started TNFi, while GPT-4 remained the most robust overall in capturing the reasons for switching—particularly when clinical documentation was sparse. Models like Llama-3-8B-Instruct and Starling-7B-beta showed relatively high concordance rates with GPT-4, suggesting these models can approximate GPT-4’s capabilities for identifying new TNFi orders. Still, GPT-4’s consistency across all facets of extraction underscores the potential advantage of larger, more general-purpose models when complete accuracy— especially in nuanced fields such as clinical pharmacovigilance—is needed.

These findings point to practical uses of LLMs in clinical care and research. First, patients with chronic inflammatory conditions, such as RA or IBD, often accumulate lengthy and intricate treatment histories over many years, making it challenging for clinicians to quickly review prior medication trials, side effects, and other complex factors that led to changes in therapy. By harnessing LLMs to summarize historical treatment notes, providers can more rapidly glean why prior regimens were discontinued—allowing for more informed decision-making and improved continuity of care. Second, enabling scalable and automated extraction of real-world reasons for treatment changes can expand the research utility of clinical notes, supporting investigations into health services challenges like insurance approvals, adverse drug reactions, and the emergence of anti-drug antibodies. Finally, with the rapid expansion of biosimilars in TNFi therapy, LLM-based systems could serve as a cornerstone of modern pharmacovigilance, proactively identifying patterns of drug resistance, adverse effects, or cost barriers. By unlocking unstructured data at scale, these approaches may inform more personalized treatment strategies and facilitate ongoing surveillance of newer therapeutics.

This study has several strengths. First, it demonstrates that GPT-4 and some open-source LLMs can extract clinically meaningful treatment information with high concordance. Second, our findings suggest that LLM-extracted insights may provide richer contextual information compared to structured medication data alone, particularly regarding reasons for TNFi switching. Third, the comparative analysis of open-source LLMs provides valuable, detailed insights into their performance relative to GPT-4, showing that some models achieve comparable accuracy for this specific task at a much lower computational cost.

Our study also has limitations. Firstly, our approach does not capture changes in medication doses, which may be relevant for treatment modifications. It also does not capture discontinuations or patients lost to follow-up. Additionally, because we only used the first clinical note associated with a switch, we may have missed relevant contextual details in some cases, or some situations in which more than one reason is discussed. In other cases, documentation of some visits may not include reasons for switching anywhere. However, given that the proportion of “other” reasons for switching was low and we performed manual chart review, the risk of missing key information is likely low. Another limitation is that our study did not explore disease-specific switching patterns in depth, nor did it account for changes to or from medications other than TNFi. Future research could refine LLM approaches to capture a broader range of unstructured data, and with-it treatment transitions. Structured medication order data are also not always reflective of medication usage and more reliable information could come from patient reported data or only relying on medication administration data.

Finally, while we compared multiple LLMs, their performance was benchmarked primarily against GPT-4 rather than expert annotations. While GPT-4 is the state of the art in LLMs, expert annotations could provide a more definitive assessment of their accuracy, since GPT-4 extracted treatment information from notes is often poorly aligned with structured medical data around medication switching, particularly medication stopping. Given the prohibitive cost of large-scale manual chart review, a combination of both evaluation methods may be the best course.^19,20^

Despite these limitations, this study highlights the potential for LLMs to extract complex treatment patterns from clinical notes at scale. As LLMs continue to improve, their integration into clinical research workflows could enhance real-world evidence generation, informing both personalized treatment strategies and healthcare policy decisions. Future studies should explore the generalizability of these methods across other institutions and treatment classes to further validate their clinical utility.

## Author Contributions

Drs Miao, Binvignat, Garcia-Agundez had full access to all of the data in the study and take responsibility for the integrity of the data and the accuracy of the data analysis.

Concept and design: Miao, Binvignat, Butte, Schmajuk, Yazdany

Acquisition, analysis, or interpretation of data: Miao, Binvignat, Garcia-Agundez, Bravo, Williams, Miao, Alaa, Rupdrapatna, Butte, Schmajuk, Yazdany

Drafting of the manuscript: Miao, Binvignat, Garcia-Agundez, Schmajuk, Yazdany Critical review of the manuscript for important intellectual content: All authors.

Statistical analysis: Miao, Binvignat, Garcia-Agundez.

Administrative, technical, or material support: Miao, Binvignat, Garcia-Agundez, Butte. Supervision: Butte, Schmajuk Yazdany

## Data Sharing Statement

Data available: No

