## Supplements for "Extracting TNFi Switching Reasons and Trajectories From Real-World Data Using Large Language Models"

### Supplemental

#### Figure S1. Clinical annotation guidelines for gold-standard labels of TNFi switching.

#### Figure S2 Average Win Rates of Large Language Models Compared to Clinical Annotation in Extracting TNFi Switching Information.

#### Table S1. TNF inhibitor drug generic and brand names.

#### Table S2. Prompts tested using automated evaluation.

#### Table S3. Examples of reasons extracted with GPT-4 from TNFi switching notes.

#### Table S4. Open-source model information.

#### Table S5. Top 25 TNFi switch pattern trajectories.

#### Table S6. Open-source language model performance on validation set compared to silver-standard labels derived from medication order data.

#### Table S7. Open-source language model performance on validation set compared to human gold-standard annotations.

#### Table S8. Reasons for treatment switching extracted using GPT-4 across prompt development and test set (n=3104).

#### Table S9. Concordance of open-source language models with GPT-4 values.

#### Figure S1. Clinical annotation guidelines for gold-standard labels of TNFi switching.

**TNFi Switching - Annotation Guidelines**

**Data overview**

In the Box Folder, you will find two CSV files

1. Annotation_set.csv
2. Med_mapping_dictionary.csv

The **Annotation_set.csv file** is where you will record your annotations, and contains the following columns. The columns highlighted in green are ones for you to fill out.

| **Column** | **Description** |
| --- | --- |
| **note_deid_note_key** | A unique identifier for each note |
| **note_text** | The contents of the note |
| **TNFi started** | Needs annotation |
| **TNFi stopped** | Needs annotation |
| **Reason type for stopping** | Needs annotation |
| **Comment** | Optional annotation |

**Annotation overview**

Here are descriptions of the columns requiring annotations. Expected values for each column are highlighted in pink (TNFi started/stopped) or blue (Reason type for stopping). Items highlighted in green indicate additional comments that should be added to the “comment” column in certain cases.

Task: Tumor necrosis factor inhibitors (TNFis) describe biologic drugs targeting TNF proteins. Using the clinical note provided, extract the following information into this JSON format: {"new_TNFi":"What new TNFi was prescribed or started? If the patient is not starting a new TNFi, write "NA"","last_TNFi":"What was the last TNFi the patient used? If none, write "NA"","reason_type_last_TNFi_stopped":"Which best describes why the last TNFi was stopped or planned to be stopped? ","full_reason_last_TNFi_stopped":"Provide a description for why the last TNFi was stopped or planned to be stopped?"} Answer:

**TNFi started/stopped**

Using information from the **note_text**, identify which contraceptive class the patient started and which one they stopped, if applicable. Each entry in these columns should be **one of the following TNFi or TNFi biosimilar values**:

- "certolizumab"
- "etanercept"
  - "etanercept-ykro"
  - "etanercept-szzs"
- "adalimumab"
  - "adalimumab-aaty"
  - "adalimumab-aacf"
  - "adalimumab-aqvh"
  - "adalimumab-fkjp”
  - "adalimumab-afzb"
  - "adalimumab-bwwd"
  - "adalimumab-adaz"
  - "adalimumab-adbm"
  - "adalimumab-atto"
- "Infliximab"
  - "infliximab-axxq"
  - "infliximab-qbtx"
  - "infliximab-abda"
  - "infliximab-dyyb"
- "golimumab"

If needed, you can use the **“med_mapping_table.csv” file** to map brand or generic names found in the note text to the contraceptive class.

- If no mapping exists, or you are not sure, please write the exact name of the TNFi **as written in the note**
- There may be redacted contraceptives (which are denoted by asterisks *****). If you are able to infer the type of TNF, feel free to write your best guess, but please add “Redacted Medication Start” AND/OR “Redacted Medication Stop” in the “Additional comments” column.
- If no contraceptive was stopped or started, write “NA” or leave the row blank.

**Reason for stopping**

Using information from the **note_text** columns, determine the reason for stopping.

- Annotate this column with one of the following reasons: "Adverse event", "Drug resistance", "Insurance/Cost","Lack of efficacy","Patient preference","Other", "NA"
  - If no reason is provided in the clinical note, write “NA”
  - If multiple reasons are available, write out additional reasons in the “Additional comments” column as a list separated by semicolons (;)
- If the reason is redacted, write your best guess if possible (otherwise leave blank) and annotate the “Additional comments” as “Redacted”
- If the reason is ambiguous, write your best guess if possible (otherwise leave blank) and annotate the “Additional comments” as “Ambiguous”

#### Figure S2 Average Win Rates of Large Language Models Compared to Clinical Annotation in Extracting TNFi Switching Information. The heatmap contains average pairwise win rates for each proprietary and open-source model evaluated on TNFi initiation and cessation data extraction. Win rates represent the proportion of instances where model A correctly identifies information while model B does not, benchmarked against gold-standard clinical annotations. Win rates for GPT-4-turbo-128k, the only proprietary model assessed, are highlighted. As an example, GPT-4-turbo-128 beats llama-3-8b-chat-hf 7% of the time (first row, second column), while the opposite happens 9% of the time (second row, first column).

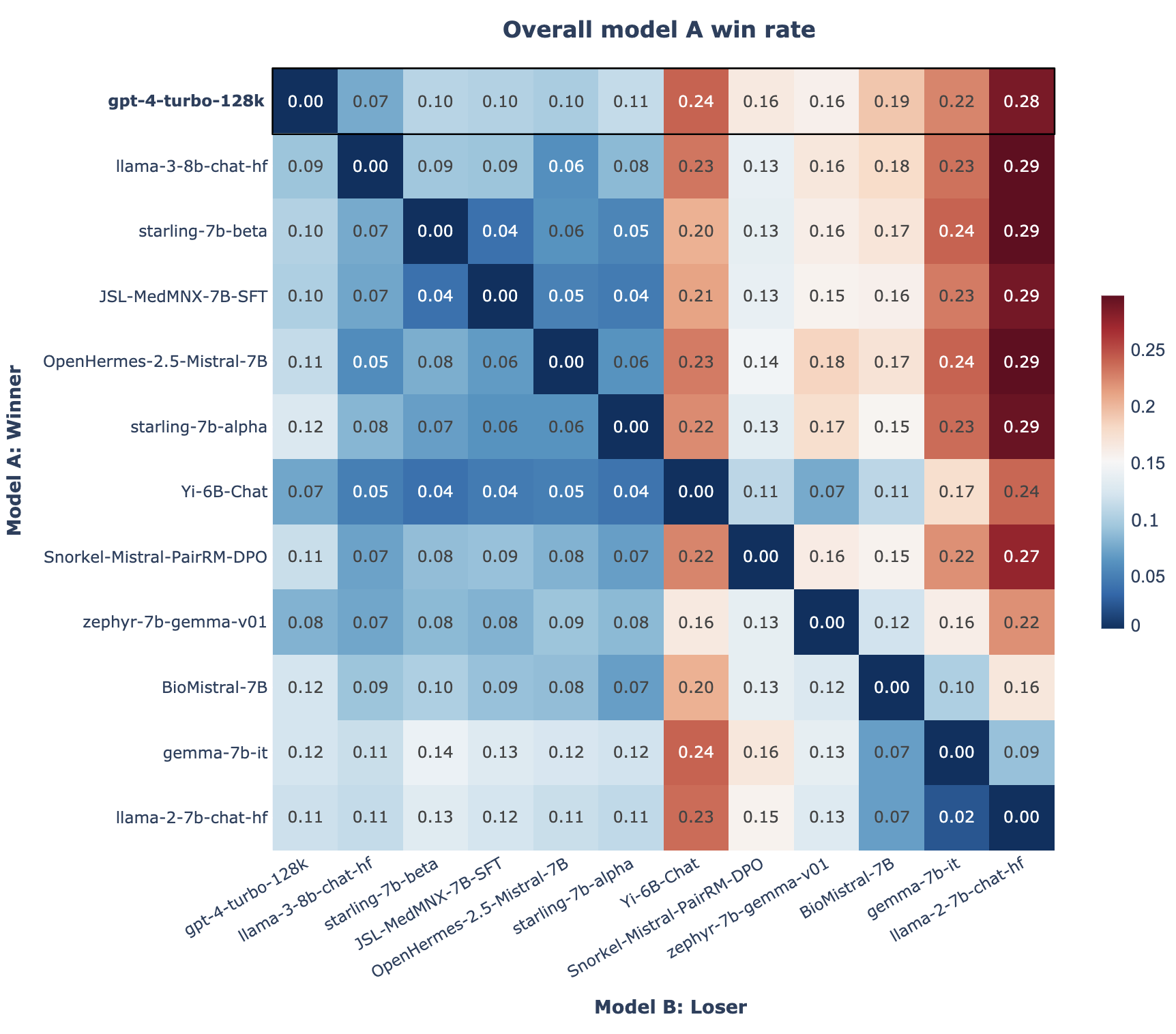

#### Table S1. TNF inhibitor drug generic and brand names.

| **Brand name** | **Generic name** |
| --- | --- |
| cimzia | certolizumab |
| enbrel | etanercept |
| humira | adalimumab |
| remicade | infliximab |
| simponi | golimumab |
| eticovo | etanercept-ykro |
| erelzi | etanercept-szzs |
| yuflyma | adalimumab-aaty |
| idacio | adalimumab-aacf |
| yusimry | adalimumab-aqvh |
| hulio | adalimumab-fkjp |
| abrilada | adalimumab-afzb |
| hadlima | adalimumab-bwwd |
| hyrimoz | adalimumab-adaz |
| cyltezo | adalimumab-adbm |
| amjevita | adalimumab-atto |
| avsola | infliximab-axxq |
| ixifi | infliximab-qbtx |
| renflexis | infliximab-abda |
| inflectra | infliximab-dyyb |

#### Table S2. Prompts tested using automated evaluation.

| **Prompt** | **Prompt Text** |
| --- | --- |
| Default | Task: Using the clinical note provided, answer the following questions - 1. What new TNF inhibitor (TNFi) biologic drug was prescribed or started? If the patient is not starting a new TNF inhibitor drug, write "NA" 2. What was the last TNF inhibitor drug the patient used? If none, write "NA" 3. Why was the last TNF inhibitor drug stopped or planned to be stopped? If no reason was provided or no TNFi was stopped, write "NA". Use the following format: {"new_TNFi":str, "last_TNFi":str, "reason_last_TNFi_stopped":str} Answer: |
| Drugs provided | Task: Cimzia (certolizumab), Enbrel (etanercept), Humira (adalimumab), Remicade (infliximab), Simponi (golimumab), Eticovo (etanercept-ykro), Erelzi (etanercept-szzs), Yuflyma (adalimumab-aaty), Idacio (adalimumab-aacf), Yusimry (adalimumab-aqvh), Hulio (adalimumab-fkjp), Abrilada (adalimumab-afzb), Hadlima (adalimumab-bwwd), Hyrimoz (adalimumab-adaz), Cyltezo (adalimumab-adbm), Amjevita (adalimumab-atto), Avsola (infliximab-axxq), Ixifi (infliximab-qbtx), Renflexis (infliximab-abda), Inflectra (infliximab-dyyb), and Renflixis (infliximab-abda) are tumor necrosis factor inihibitor (TNFi) biologic drugs. Using the clinical note provided, extract the following information into this JSON format: {"new_TNFi":"What new TNFi was prescribed or started? If the patient is not starting a new TNFi, write "NA"","last_TNFi":"What was the last TNFi the patient used? If none, write "NA"","reason_last_TNFi_stopped":"Why was the last TNFi stopped or planned to be stopped? If no reason was provided or no TNFi was stopped, write "NA""} Answer: |
| Reasons provided | Task: Tumor necrosis factor inhibitors (TNFis) describe biologic drugs targeting TNF proteins. Using the clinical note provided, extract the following information into this JSON format: {"new_TNFi":"What new TNFi was prescribed or started? If the patient is not starting a new TNFi, write "NA"","last_TNFi":"What was the last TNFi the patient used? If none, write "NA"","Reason for Switching":"Which best describes why the last TNFi was stopped or planned to be stopped? "Adverse event", "Drug resistance", "Insurance/Cost","Lack of efficacy","Patient preference","Other", "NA"","full_reason_last_TNFi_stopped":"Provide a description for why the last TNFi was stopped or planned to be stopped?"} Answer: |
| All values provided | Task: Cimzia (certolizumab), Enbrel (etanercept), Humira (adalimumab), Remicade (infliximab), Simponi (golimumab), Eticovo (etanercept-ykro), Erelzi (etanercept-szzs), Yuflyma (adalimumab-aaty), Idacio (adalimumab-aacf), Yusimry (adalimumab-aqvh), Hulio (adalimumab-fkjp), Abrilada (adalimumab-afzb), Hadlima (adalimumab-bwwd), Hyrimoz (adalimumab-adaz), Cyltezo (adalimumab-adbm), Amjevita (adalimumab-atto), Avsola (infliximab-axxq), Ixifi (infliximab-qbtx), Renflexis (infliximab-abda), Inflectra (infliximab-dyyb), and Renflixis (infliximab-abda) are tumor necrosis factor inihibitor (TNFi) biologic drugs. Using the clinical note provided, extract the following information into this JSON format: {"new_TNFi":"What new TNFi was prescribed or started? If the patient is not starting a new TNFi, write "NA"","last_TNFi":"What was the last TNFi the patient used? If none, write "NA"","Reason for Switching":"Which best describes why the last TNFi was stopped or planned to be stopped? "Adverse event", "Drug resistance", "Insurance/Cost","Lack of efficacy","Patient preference","Other", "NA"","full_reason_last_TNFi_stopped":"Provide a description for why the last TNFi was stopped or planned to be stopped?"} Answer: |

#### Table S3. Examples of reasons extracted with GPT-4 from TNFi switching notes. Notes have been de identified with filter that may in some cases redacted some medical informations misidentified as PHI (Private Health Information)

| **Note excerpt** | **GPT-4 extracted TNFi started** | **GPT-4 extracted TNFi stopped** | **GPT-4 extracted reason** |
| --- | --- | --- | --- |
| Biologic medication authorization request Diagnosis (x) Ulcerative Colitis (x) Pancolitis ( ) left sided ( ) rectosigmoid ICD ***** ***** *****.011 ( x ) new therapy ( ) ongoing therapy Patient pertinent history ( please provide as much detail as possible including reason to start biologic, reason for specific dose): infantile onset ulcerative colitis, initially well-controlled with infliximab 10mg/kg monthly, but with loss of response as evidenced by active colitis on endoscopy with adequate infliximab level and no antibody formation; transitioned to Entyvio 300mg monthly for the past 6 months, unable to wean steroids without disease relapse, also receiving subcutaneous methotrexate. Therefore requesting transition to adalimumab.  *[392 tokens omitted for length]* | adalimumab | infliximab | Lack of efficacy |
| *[2066 tokens omitted for length]*  At her last visit, we tried escalating her therapy to infliximab (she had a strong preference for this medication) and starting *****, but she unfortunately stopped her ***** and had an infusion reaction to infliximab.  *[215 tokens omitted for length]*  --stop infliximab/***** --start plaquenil 200mg, will refer for baseline eye exam --continue prednisone 5mg --will start cimzia, hopefully this will not result in antibody formation as it is less immunogenic than infliximab, and she feels that she responded to infliximab in the past  *[264 tokens omitted for length]* | cimzia | infliximab | Adverse event |
| Prior Authorization: adalimumab 40mg/wk subcutaneous Diagnosis Code: *****.2 Hidradenitis suppurativa Previous failed alternative therapies: Colchicine - intolerable GI side effects Dapsone - ineffective etanercept - no improvement Infliximab - excellent control of her hidradenitis but infusions too expensive for longterm  *[89 tokens omitted for length]*  Medical justification: patient responds well to *****/infliximab for her HS. Adalimumab is FDA approved for her HS and should give similar efficacy without expensive monthly infusions | adalimumab | infliximab | Insurance/Cost |
| Pt with anti-drug antibodies to adalimumab >100 And low adalimumab level 0.9 This is likely related to pts report of decreased benefit of meds for RA joint pain and swelling. Would like to switch pt to etanercept. ***** submit for prior auth. | etanercept | adalimumab | Drug Resistance |
| *[80 tokens omitted for length]*  *After the initial 2 doses of Humira, he was not able to access further doses due to unacceptable co-pay and reimbursement mechanism. Despite this, overall he feels that he has done quite well. He has mostly remained on prednisone 5mg since our last visit.*  *[500 tokens omitted for length]*  *Initially on Enbrel (*****-*****), then Humira (*****-*****), then ***** (*****-*****), feels like these would help for a short time but then he would plateau.*  *[1593 tokens omitted for length]*  After our initial evaluation, we decided to return to ***** ***** management of MTX + *****. Unfortunately having copay issues with Humira, will try Enbrel instead.  *[1133 tokens omitted for length]* | Enbrel | Humira | Insurance/Cost |

#### Table S4. Open source model information.

| **Model name** | **Base Model** | **Release date** | **Reference or model repository** |
| --- | --- | --- | --- |
| Llama-2-7b-Chat | Llama-2-7B | Jul 18, 2023 | Touvron et al, 2023^21^ |
| OpenHermes-2.5-Mistral-7B | Mistral-7B | Oct 29, 2023 | <https://huggingface.co/teknium/OpenHermes-2.5-Mistral-7B> |
| Yi-6B-Chat | Yi-6B | Nov 22, 2023 | 01.AI 2024^22^ |
| Starling-7b-alpha | Mistral-7B | Nov 25, 2023 | Zhu et al, 2023^23^ |
| Snorkel-Mistral-PairRM-DPO | Mistral-7B | Jan 19, 2024 | <https://huggingface.co/snorkelai/Snorkel-Mistral-PairRM-DPO> |
| BioMistral-7B | Mistral-7B | Feb 14, 2024 | Labrak et al, 2024^24^ |
| Gemma-7B-IT | Gemma-7B | Feb 21, 2024 | <https://huggingface.co/google/gemma-7b-it> |
| Zephyr-7b-gemma-v01 | Gemma-7B | Mar 1, 2024 | <https://huggingface.co/HuggingFaceH4/zephyr-7b-gemma-v0.1> |
| Starling-7b-beta | Mistral-7B | Mar 19, 2024 | <https://huggingface.co/Nexusflow/Starling-LM-7B-beta> |
| JSL-MedMNX-7B-SFT | Starling-LM-7B-beta & Mistral-7B-v0.1 | April 15, 2024 | <https://huggingface.co/johnsnowlabs/JSL-MedMNX-7B-SFT> |
| Llama-3-8b-Instruct | Llama-3-8B | April 18, 2024 | <https://huggingface.co/meta-llama/Meta-Llama-3-8B-Instruct> |

#### Table S5. Top 25 TNFi switch pattern trajectories.

| **First medication** | **Second medication** | **Third medication** | **Count** | **%** |
| --- | --- | --- | --- | --- |
| etanercept | adalimumab |  | 546 | 3.6% |
| adalimumab | infliximab |  | 265 | 1.7% |
| infliximab | adalimumab |  | 241 | 1.6% |
| infliximab | infliximab-dyyb |  | 159 | 1.0% |
| adalimumab | etanercept |  | 148 | 1.0% |
| adalimumab | certolizumab |  | 118 | 0.8% |
| etanercept | adalimumab | etanercept | 71 | 0.5% |
| infliximab | adalimumab | etanercept | 71 | 0.5% |
| adalimumab | infliximab | infliximab-dyyb | 64 | 0.4% |
| infliximab-dyyb | infliximab | infliximab-dyyb | 62 | 0.4% |
| infliximab-dyyb | infliximab |  | 62 | 0.4% |
| infliximab | infliximab-dyyb | infliximab | 56 | 0.4% |
| adalimumab | golimumab |  | 54 | 0.4% |
| infliximab | adalimumab | infliximab | 54 | 0.4% |
| etanercept | adalimumab | infliximab | 54 | 0.4% |
| etanercept | infliximab | infliximab-dyyb | 48 | 0.3% |
| etanercept | infliximab |  | 48 | 0.3% |
| infliximab-dyyb | infliximab | adalimumab | 47 | 0.3% |
| etanercept | infliximab | adalimumab | 47 | 0.3% |
| adalimumab | infliximab | adalimumab | 47 | 0.3% |
| etanercept | certolizumab |  | 39 | 0.3% |
| etanercept | adalimumab | certolizumab | 38 | 0.3% |
| infliximab | adalimumab | certolizumab | 38 | 0.3% |
| adalimumab | infliximab-dyyb |  | 37 | 0.2% |
| adalimumab | infliximab-dyyb | infliximab | 37 | 0.2% |

#### Table S6. Open-source language model performance on validation set compared to silver-standard labels derived from medication order data.

| **Model** | **Prompt** | **Extraction Task** | **All values (microF1)** | **Null values dropped (microF1)** | **Non-null values (n)** |
| --- | --- | --- | --- | --- | --- |
| gpt-4-turbo-128k | default-task | TNFi Stopped | 0.41 | 0.61 | 76 |
| gpt-4-turbo-128k | default-task | New TNFi Started | 0.46 | 0.88 | 52 |
| gpt-4-turbo-128k | all-values-provided | TNFi Stopped | 0.41 | 0.59 | 78 |
| gpt-4-turbo-128k | all-values-provided | New TNFi Started | 0.46 | 0.90 | 50 |
| gpt-4-turbo-128k | drugs-provided | TNFi Stopped | 0.38 | 0.54 | 79 |
| gpt-4-turbo-128k | drugs-provided | New TNFi Started | 0.47 | 0.90 | 52 |
| gpt-4-turbo-128k | reasons-provided | TNFi Stopped | 0.41 | 0.63 | 71 |
| gpt-4-turbo-128k | reasons-provided | New TNFi Started | 0.50 | 0.89 | 56 |
| starling-7b-beta | reasons-provided | TNFi Stopped | 0.43 | 0.83 | 52 |
| starling-7b-beta | reasons-provided | New TNFi Started | 0.60 | 0.88 | 76 |
| llama-3-8b-chat-hf | reasons-provided | TNFi Stopped | 0.44 | 0.79 | 56 |
| llama-3-8b-chat-hf | reasons-provided | New TNFi Started | 0.57 | 0.83 | 76 |
| JSL-MedMNX-7B-SFT | reasons-provided | TNFi Stopped | 0.41 | 0.82 | 49 |
| JSL-MedMNX-7B-SFT | reasons-provided | New TNFi Started | 0.59 | 0.83 | 81 |
| OpenHermes-2.5-Mistral-7B | reasons-provided | TNFi Stopped | 0.38 | 0.88 | 42 |
| OpenHermes-2.5-Mistral-7B | reasons-provided | New TNFi Started | 0.61 | 0.82 | 85 |
| starling-7b-alpha | reasons-provided | TNFi Stopped | 0.37 | 0.87 | 39 |
| starling-7b-alpha | reasons-provided | New TNFi Started | 0.60 | 0.82 | 84 |
| Yi-6B-Chat | reasons-provided | TNFi Stopped | 0.39 | 0.63 | 67 |
| Yi-6B-Chat | reasons-provided | New TNFi Started | 0.57 | 0.83 | 77 |
| Snorkel-Mistral-PairRM-DPO | reasons-provided | TNFi Stopped | 0.40 | 0.76 | 51 |
| Snorkel-Mistral-PairRM-DPO | reasons-provided | New TNFi Started | 0.54 | 0.77 | 79 |
| zephyr-7b-gemma-v01 | reasons-provided | TNFi Stopped | 0.34 | 0.56 | 64 |
| zephyr-7b-gemma-v01 | reasons-provided | New TNFi Started | 0.46 | 0.94 | 48 |
| BioMistral-7B | reasons-provided | TNFi Stopped | 0.22 | 0.86 | 21 |
| BioMistral-7B | reasons-provided | New TNFi Started | 0.38 | 0.80 | 45 |
| gemma-7b-it | reasons-provided | TNFi Stopped | 0.03 | 1.00 | 2 |
| gemma-7b-it | reasons-provided | New TNFi Started | 0.05 | 0.89 | 5 |
| llama-2-7b-chat-hf | reasons-provided | TNFi Stopped | 0.04 | 0.22 | 14 |
| llama-2-7b-chat-hf | reasons-provided | New TNFi Started | 0.10 | 0.62 | 13 |

#### Table S7. Open-source language model performance on validation set compared to human gold-standard annotations.

| **Extraction Task** | **Model** | **Prompt** | **microF1** |
| --- | --- | --- | --- |
| **TNFi Stopped** | **starling-7b-alpha** | **reasons-provided** | **0.897** |
|  | starling-7b-beta | reasons-provided | 0.877 |
|  | JSL-MedMNX-7B-SFT | reasons-provided | 0.852 |
|  | OpenHermes-2.5-Mistral-7B | reasons-provided | 0.836 |
|  | llama-3-8b-chat-hf | reasons-provided | 0.818 |
|  | Snorkel-Mistral-PairRM-DPO | reasons-provided | 0.801 |
|  | BioMistral-7B | reasons-provided | 0.795 |
|  | gpt-4-turbo-128k | reasons-provided | 0.749 |
|  | gemma-7b-it | reasons-provided | 0.733 |
|  | gpt-4-turbo-128k | default-task | 0.715 |
|  | zephyr-7b-gemma-v01 | reasons-provided | 0.699 |
|  | gpt-4-turbo-128k | all-values-provided | 0.680 |
|  | gpt-4-turbo-128k | drugs-provided | 0.664 |
|  | Yi-6B-Chat | reasons-provided | 0.660 |
|  | llama-2-7b-chat-hf | reasons-provided | 0.644 |
| **TNFi Started** | **gpt-4-turbo-128k** | **all-values-provided** | **0.801** |
|  | gpt-4-turbo-128k | drugs-provided | 0.788 |
|  | starling-7b-beta | reasons-provided | 0.788 |
|  | gpt-4-turbo-128k | reasons-provided | 0.774 |
|  | JSL-MedMNX-7B-SFT | reasons-provided | 0.770 |
|  | gpt-4-turbo-128k | default-task | 0.760 |
|  | zephyr-7b-gemma-v01 | reasons-provided | 0.760 |
|  | starling-7b-alpha | reasons-provided | 0.753 |
|  | OpenHermes-2.5-Mistral-7B | reasons-provided | 0.753 |
|  | llama-3-8b-chat-hf | reasons-provided | 0.753 |
|  | Snorkel-Mistral-PairRM-DPO | reasons-provided | 0.719 |
|  | Yi-6B-Chat | reasons-provided | 0.719 |
|  | BioMistral-7B | reasons-provided | 0.692 |
|  | gemma-7b-it | reasons-provided | 0.603 |
|  | llama-2-7b-chat-hf | reasons-provided | 0.575 |
| **Reason for Switching** | **gpt-4-turbo-128k** | **all-values-provided** | **0.825** |
|  | llama-3-8b-chat-hf | reasons-provided | 0.823 |
|  | gpt-4-turbo-128k | reasons-provided | 0.817 |
|  | OpenHermes-2.5-Mistral-7B | reasons-provided | 0.779 |
|  | JSL-MedMNX-7B-SFT | reasons-provided | 0.706 |
|  | starling-7b-alpha | reasons-provided | 0.704 |
|  | gemma-7b-it | reasons-provided | 0.680 |
|  | zephyr-7b-gemma-v01 | reasons-provided | 0.659 |
|  | starling-7b-beta | reasons-provided | 0.638 |
|  | Snorkel-Mistral-PairRM-DPO | reasons-provided | 0.635 |
|  | BioMistral-7B | reasons-provided | 0.623 |
|  | llama-2-7b-chat-hf | reasons-provided | 0.601 |
|  | Yi-6B-Chat | reasons-provided | 0.468 |

#### Table S8. Reasons for treatment switching extracted using GPT-4 across prompt development and test set (n=3104).

| **Reason type** | **Count** | **Proportion** |
| --- | --- | --- |
| NA/Unknown | 2106 | - |
| Lack of efficacy | 568 | 56.9% |
| Adverse event | 135 | 13.5% |
| Insurance/Cost | 108 | 10.8% |
| Patient preference | 82 | 8.2% |
| Other | 54 | 5.4% |
| Drug resistance | 51 | 5.1% |

#### Table S9. Concordance of open-source language models with GPT-4 values.

|  | **Previous TNFi** | **TNFi Started** | **Reason Stopped** | **Mean** | **SD** |
| --- | --- | --- | --- | --- | --- |
| gpt-4-turbo-128k | 100.0% | 100.0% | 100.0% | 100.0% | 0.0% |
| llama-3-8b-chat-hf | 82.9% | 81.5% | 82.9% | 82.4% | 0.6% |
| starling-7b-beta | 80.8% | 81.5% | 68.5% | 76.9% | 6.0% |
| JSL-MedMNX-7B-SFT | 78.1% | 80.1% | 69.9% | 76.0% | 4.4% |
| OpenHermes-2.5-Mistral-7B | 72.6% | 75.3% | 81.5% | 76.5% | 3.7% |
| starling-7b-alpha | 72.6% | 76.7% | 74.0% | 74.4% | 1.7% |
| Yi-6B-Chat | 68.5% | 75.3% | 45.9% | 63.2% | 12.6% |
| Snorkel-Mistral-PairRM-DPO | 73.3% | 69.9% | 64.4% | 69.2% | 3.7% |
| zephyr-7b-gemma-v01 | 71.9% | 83.6% | 65.1% | 73.5% | 7.6% |
| BioMistral-7B | 59.6% | 73.3% | 61.0% | 64.6% | 6.2% |
| gemma-7b-it | 52.7% | 65.1% | 66.4% | 61.4% | 6.2% |
| llama-2-7b-chat-hf | 44.5% | 61.0% | 60.3% | 55.3% | 7.6% |
